# Methods for Estimation of SARS-CoV-2 Seroprevalence and Reported COVID-19 Cases in U.S. Children, August 2020—May 2021

**DOI:** 10.1101/2021.09.26.21263756

**Authors:** Alexia Couture, Casey Lyons, Megha L. Mehrotra, Lynn Sosa, Ngozi Ezike, Catherine M. Brown, Stephanie Yendell, Ihsan A. Azzam, Božena J Katić, Anna Cope, Kristen Dickerson, John Dunn, L. Brannon Traxler, Lora B. Davis, Farah S. Ahmed, Jolianne Stone, Carrie Reed, Kristie E. N. Clarke, Brendan Flannery, Myrna D. Charles

## Abstract

**Background and Objectives:** Case-based surveillance of pediatric COVID-19 cases underestimates the prevalence of SARS-CoV-2 infections among children and adolescents. Our objectives were to: 1) estimate monthly SARS-CoV-2 antibody seroprevalence among children aged 0-17 years and 2) calculate ratios of SARS-CoV-2 infections to reported COVID-19 cases among children and adolescents in 14 U.S. states.

**Methods:** Using data from commercial laboratory seroprevalence surveys, we estimated monthly SARS-CoV-2 antibody seroprevalence among children aged 0-17 years from August 2020 through May 2021. Seroprevalence estimates were based on SARS-CoV-2 anti-nucleocapsid immunoassays from February to May 2021. We compared estimated numbers of children infected with SARS-CoV-2 by May 2021 to cumulative incidence of confirmed and probable COVID-19 cases from case-based surveillance, and calculated infection: case ratios by state and type of anti-SARS-CoV-2 nucleocapsid immunoassay used for seroprevalence testing.

**Results:** Analyses included 67,321 serum specimens tested for SARS-CoV-2 antibodies among children in 14 U.S. states. Estimated ratios of SARS-CoV-2 infections to reported confirmed and probable COVID-19 cases among children and adolescents varied by state and type of immunoassay, ranging from 0.8-13.3 in May 2021.

**Conclusions:** Through May 2021, the majority of children in selected states did not have detectable SARS-CoV-2 nucleocapsid antibodies. Case-based surveillance underestimated the number of children infected with SARS-CoV-2, however the predicted extent of the underestimate varied by state, immunoassay, and over time. Continued monitoring of pediatric SARS-CoV-2 antibody seroprevalence should inform prevention and vaccination strategies.

## Background

Since July 2020, CDC has partnered with commercial laboratories to conduct repeated cross-sectional seroprevalence studies using residual clinical serum specimens to test for the presence of SARS-CoV-2 antibodies.^1, 2^ In unvaccinated individuals, detection of specific antibodies to SARS-CoV-2 proteins in serum indicates previous infection.^3, 4^ Laboratory assays that detect nucleocapsid (N) antibodies identify individuals with a history of SARS-CoV-2 infection regardless of immunization status because currently approved or authorized vaccines in the U.S. elicit antibodies to SARS-CoV-2 spike protein.^2^ While commercial laboratory sero-surveillance has provided seroprevalence estimates across the age spectrum, including persons aged 0-17 years,^1^ differences in seroprevalence among pediatric age groups have not been evaluated. Projected numbers of children and adolescents with evidence of past SARS-CoV-2 infection may be contrasted with reported pediatric COVID-19 cases to estimate an infection to case ratio. Projections of SARS-CoV-2 infections among children and adolescents based on case-based surveillance may be less reliable than those for older age groups because children may be less likely to present with symptoms and be tested for SARS-CoV-2 infection. Analyses of case-surveillance data from health departments have used different case definitions and age groups for estimating COVID-19 incidence among children and adolescents.^5^ To investigate trends in estimated SARS-CoV-2 infections among children and adolescents, we analyzed commercial laboratory serosurvey data from 14 U.S. states and compared projected incidence of SARS-CoV-2 infection with case-based surveillance for confirmed and probable COVID-19 cases among pediatric age groups. This manuscript presents the methods used for this analysis and state-level trends in reported COVID-19 cases and estimated seroprevalence among children aged 0-17 years in 14 U.S. states.

## Methods

Methods for the Nationwide Commercial Laboratory Seroprevalence Survey have been previously described.^1, 2^ In brief, CDC partnered with three commercial laboratories to test a random sample of residual sera submitted for routine clinical testing for the presence of SARS-CoV-2 antibodies using one of three commercial assays authorized under EUA: Elecsys® Anti-SARS-CoV-2 Assay (N protein; Roche, Indianapolis, IN) and ARCHITECT™ SARS-CoV-2 IgG Assay (N protein; Abbott, Chicago, IL), both targeting the viral N protein, and VITROS® Anti-SARS-CoV-2 IgG Assay (spike (S) protein; Ortho-Clinical Diagnostics, Raritan, NJ) targeting the viral spike protein. Proportions of sera tested by each immunoassay varied by state and calendar month (Figure S1). All three assays were used from August 2020 to January 2021. From February to May 2021, when COVID-19 vaccination was becoming more widely available, all serum specimens from states included in this analysis were tested for either pan-immunoglobulin (Ig) (Roche Elecsys) or IgG-specific (Abbott ARCHITECT) anti-nucleocapsid antibodies. For seroprevalence estimates, we included commercial laboratory data irrespective of type of immunoassay used. We included data from states in which commercial laboratories tested ≥100 residual specimens from individuals aged 0-17 years for each month from August 2020 to May 2021.

To estimate SARS-CoV-2 seroprevalence among pediatric age groups (ages 0-4, 5-11, and 12-17 years), we calculated weighted estimates of seropositivity for each state and month of specimen collection, using de-identified patient-level data from the commercial laboratory seroprevalence survey and individual weights to account for differences in proportions of children and adolescents included in serosurveys and the state population by age group, sex, and metropolitan versus rural residence.^1^ As previously described, we estimated population-weighted seroprevalence and 95% confidence intervals (CI) accounting for sensitivity and specificity of the assays using an iterative post-stratification process known as raking and layered bootstrapping, with adjusted seroprevalence defined as the mean of the bootstrap distribution and upper and lower confidence limits set at the 2.5th and 97.5th percentiles.^6^ To estimate cumulative SARS-CoV-2 infections among persons aged 0-17 years as well as by pediatric age groups, we multiplied monthly mean seroprevalence estimates, and upper and lower confidence limits, by the population aged 0-17 years in each state for each month.^7^

For COVID-19 case-based surveillance, CDC receives individual-level data (including age, sex, dates of illness onset, and specimen collection) for probable and laboratory-confirmed COVID-19 cases from jurisdictional health departments through the COVID-19 Case Report Form and the National Notifiable Diseases Surveillance System.^8, 9^ For each reported case, date of illness was defined as either symptom onset (if reported), the earliest clinical date on case report forms, or date reported to CDC. For this analysis, we included individual-level case data from states with ≥90% concordance between daily aggregate case counts and cumulative individual case reports from March 2020 through May 2021 and with case patient age recorded for >90% of individual case reports. Data from 14 states were included in this analysis: California, Connecticut, Illinois, Kansas, Massachusetts, Minnesota, Nevada, New Jersey, North Carolina, Ohio, Oklahoma, South Carolina, Tennessee, and Washington. Connecticut contributed additional data from their state database.

To calculate ratios of infections to reported cases, we divided estimated cumulative infections (with upper and lower confidence limits) for each month by the number of confirmed and probable COVID-19 cases in persons aged 0-17 years reported by each state health department since January 2020 through the 15^th^ of each month from September 2020 through May 2021. Analyses were conducted in SAS Software, version 9.4 (SAS Institute), and R, version 3.6.3 (R Core Team).

### State-level trends in reported COVID-19 cases and estimated seroprevalence among children aged 0-17 years in 14 U.S. states

For the 14 U.S. states included in the analysis, a total of 67,321 residual serum specimens were tested from persons aged 0-17 years during August 2020 through May 2021. Proportions of pediatric specimens tested by each of the three serologic assays varied by state and calendar month (Figure S1). After January 2021, serum specimens in 8 states were tested for anti-nucleocapsid antibodies using the pan-Ig Roche Elecsys and in six states using the IgG specific Abbott ARCHITECT.

Figures S2 A through N compare projected numbers of SARS-CoV-2 infections by the end of August 2020 and May 2021 with cumulative numbers of reported COVID-19 cases (confirmed and probable) among children aged 0-17 years in each state. Estimated ratios of SARS-CoV-2 infections to reported cases among persons aged 0–17 years were higher in August 2020, ranging from 3.2 in Kansas to 58.2 in New Jersey, compared to May 2021, ranging from 0.8 in Connecticut to 8.9 in Ohio. Overall, pediatric age-specific infection:case ratios of SARS-CoV-2 infections to reported COVID-19 cases were highest during August to October 2020. From November 2020 through May 2021, infection:case ratios remained relatively stable. Higher ranges of infection:case ratios were observed for states in which commercial laboratories used the pan-Ig anti-nucleocapsid immunoassay (range, 4.7-8.9) versus for states where the IgG specific immunoassay was used (range, 0.8-3.0).

## Conclusion

Overall, these findings provide evidence of higher rates of COVID-19 among children than detected by case-based surveillance. Through May 2021, the majority of children did not have serologic evidence of past SARS-CoV-2 infection.

## Data Availability

Deidentified individual participant data will not be made available.

## Abbreviations

CDC: Centers of Disease Control and Prevention
MIS-C: Multisystem inflammatory syndrome in children
EUA: Emergency Use Authorization
FDA: U.S. Food and Drug Administration
ACIP: Advisory Committee on Immunizations Practices
N: Nucleocapsid
S: Spike
Ig: Immunoglobulin
CI: Confidence intervals

## Acknowledgments

We thank the individuals involved in COVID-19 response at state and local health departments in California, Connecticut, Illinois, Kansas, Massachusetts, Minnesota, Nevada, New Jersey, North Carolina, Ohio, Oklahoma, South Carolina, Tennessee, and Washington, employees of commercial laboratories conducting the national serosurvey, and members of the CDC COVID-19 Response, including Amitabh Suthar, Radhika Gharpure, Dawona Hough, Denise Sheriff, Stephanie Hinton, Jennifer Frazier, Rebecca Sabo, Krystal Gayle, Alicia Dunajcik, Yonathan Gebru, Neela Persad, Fija Scipio, Laura Hill, and Kia Padgett.

---

**Supplementary Figure 1:**
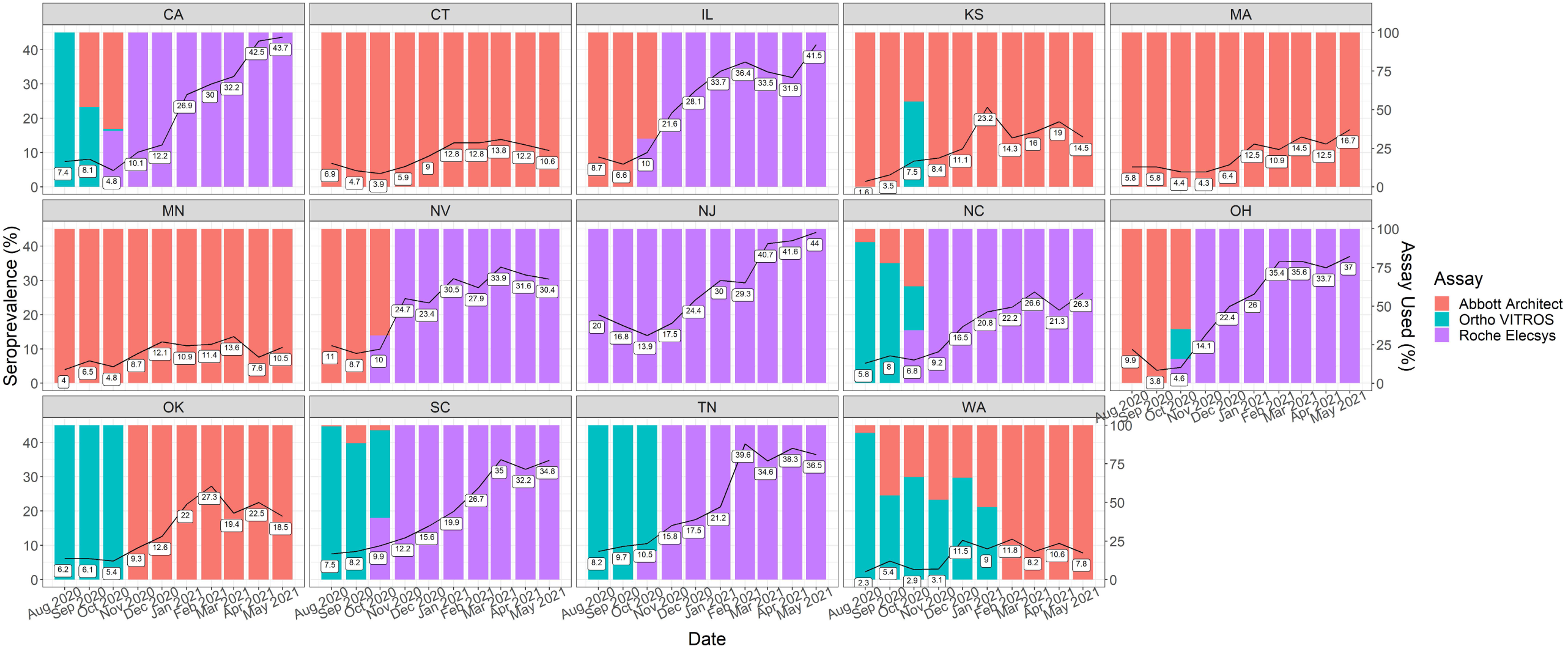
Monthly population-weighted seroprevalence among children and adolescents aged 0-17 years in 14 U.S. states included in analysis, with proportions of pediatric serum specimens tested month by each commercial immunoassay. Commercial laboratories tested randomly selected, de-duplicated residual sera for the presence of SARS-CoV-2 antibodies using one of three FDA approved commercial immunoassays under Emergency Use Authorization: Roche Elecsys Anti-SARS-CoV-2 pan-immunoglobulin (Ig) immunoassay, Abbott ARCHITECT SARS-CoV-2 IgG immunoassay and Ortho-Clinical Diagnostics VITROS SARS-CoV-2 IgG immunoassay).

**Supplementary Figure 2:**
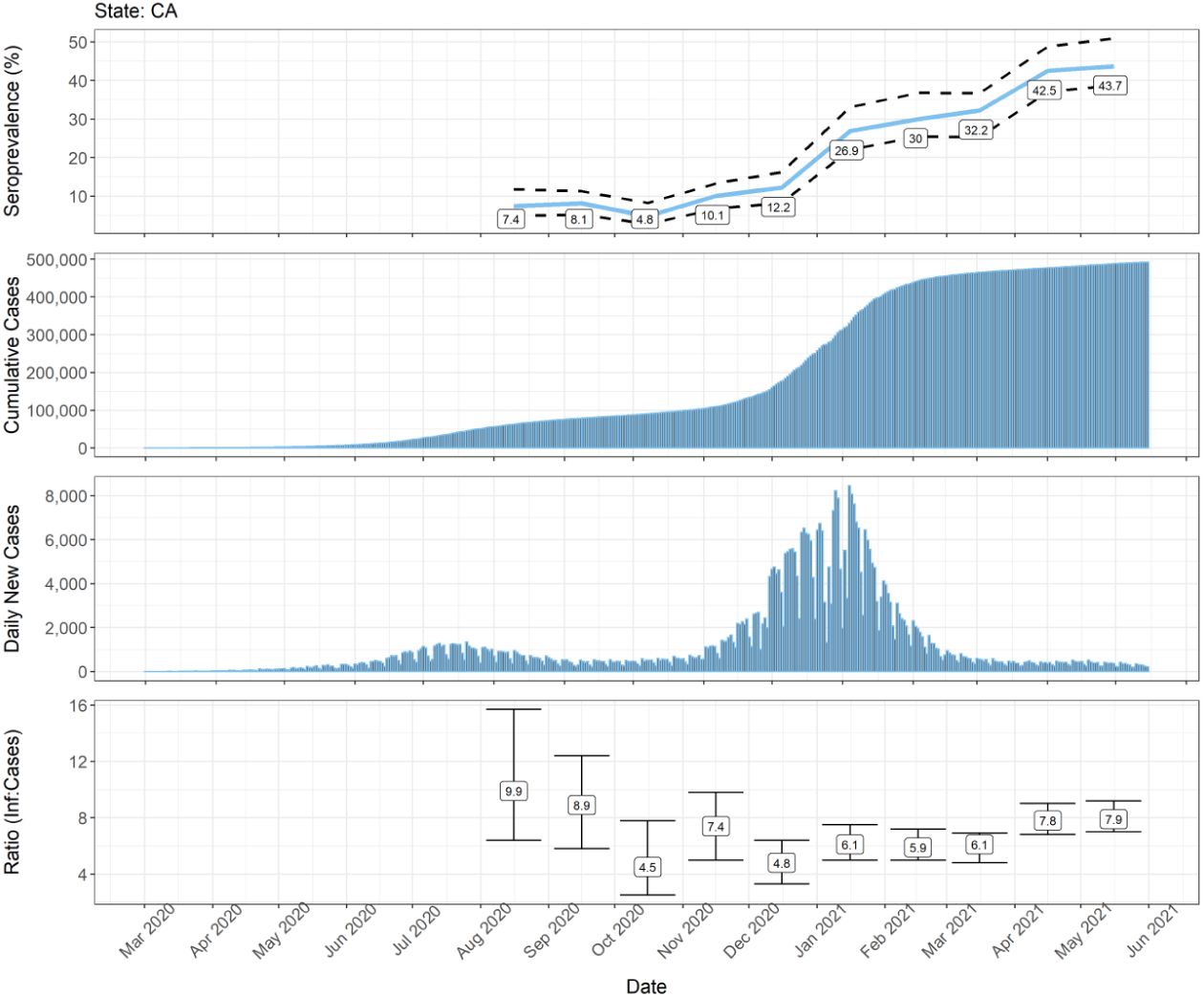
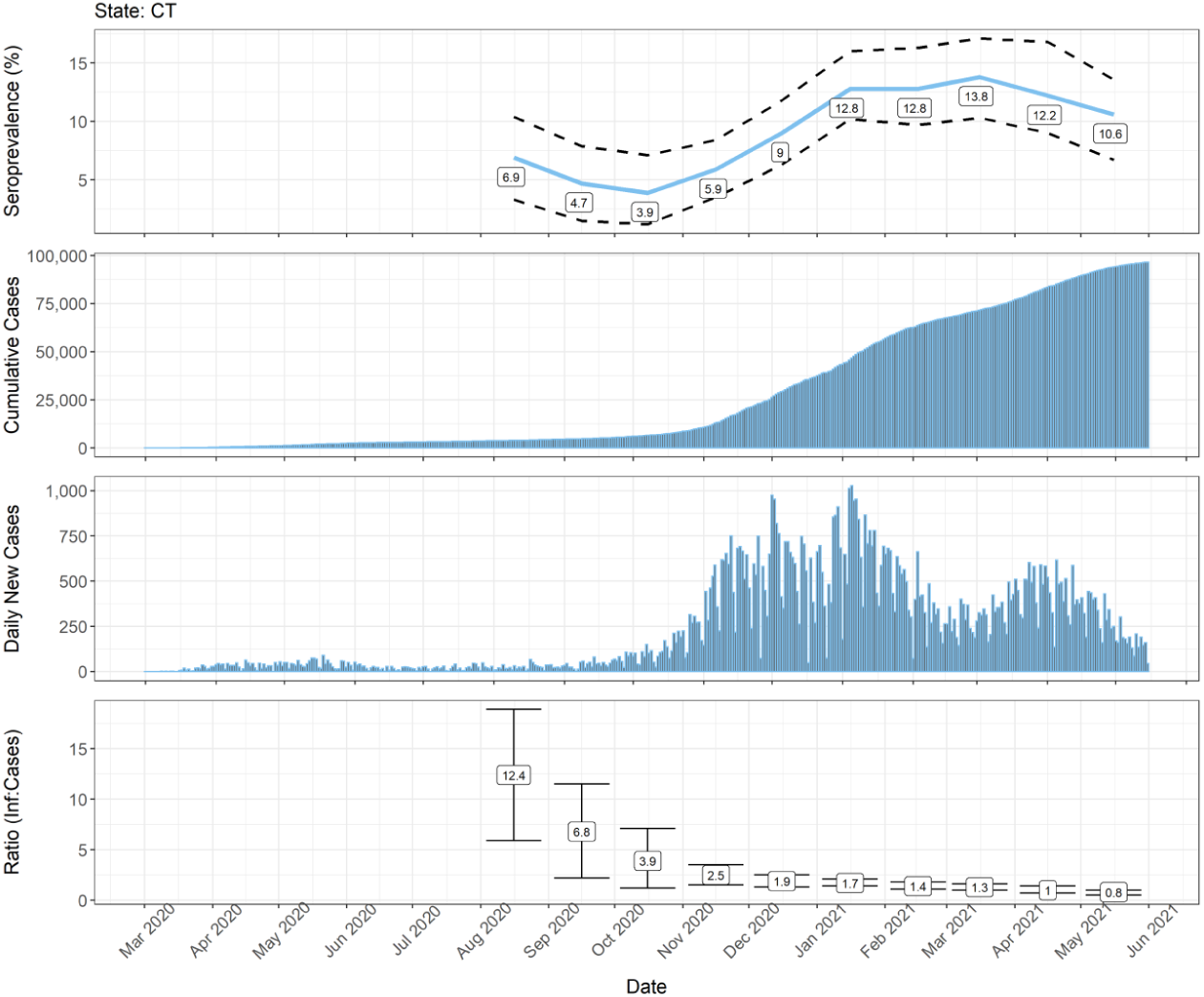
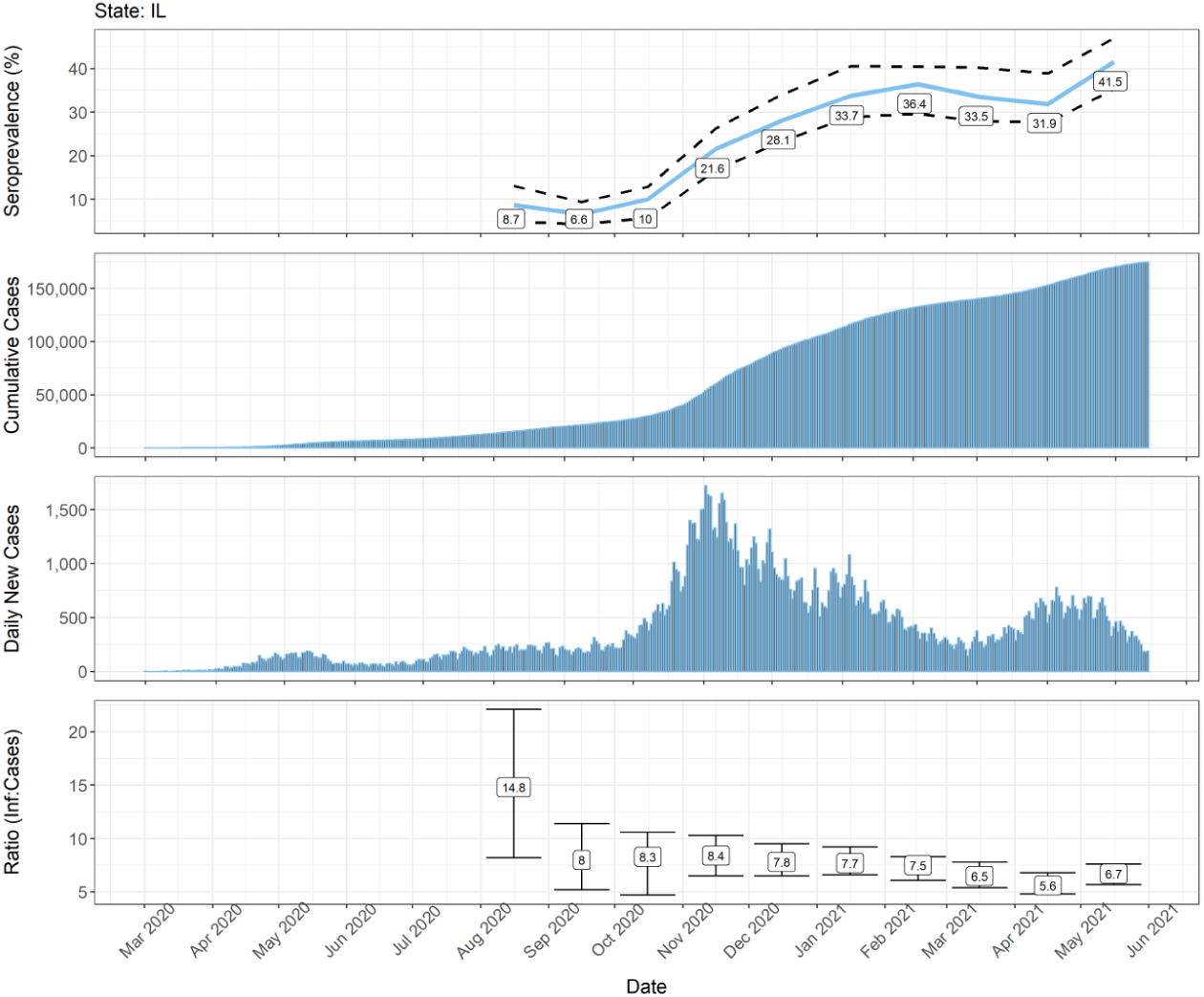
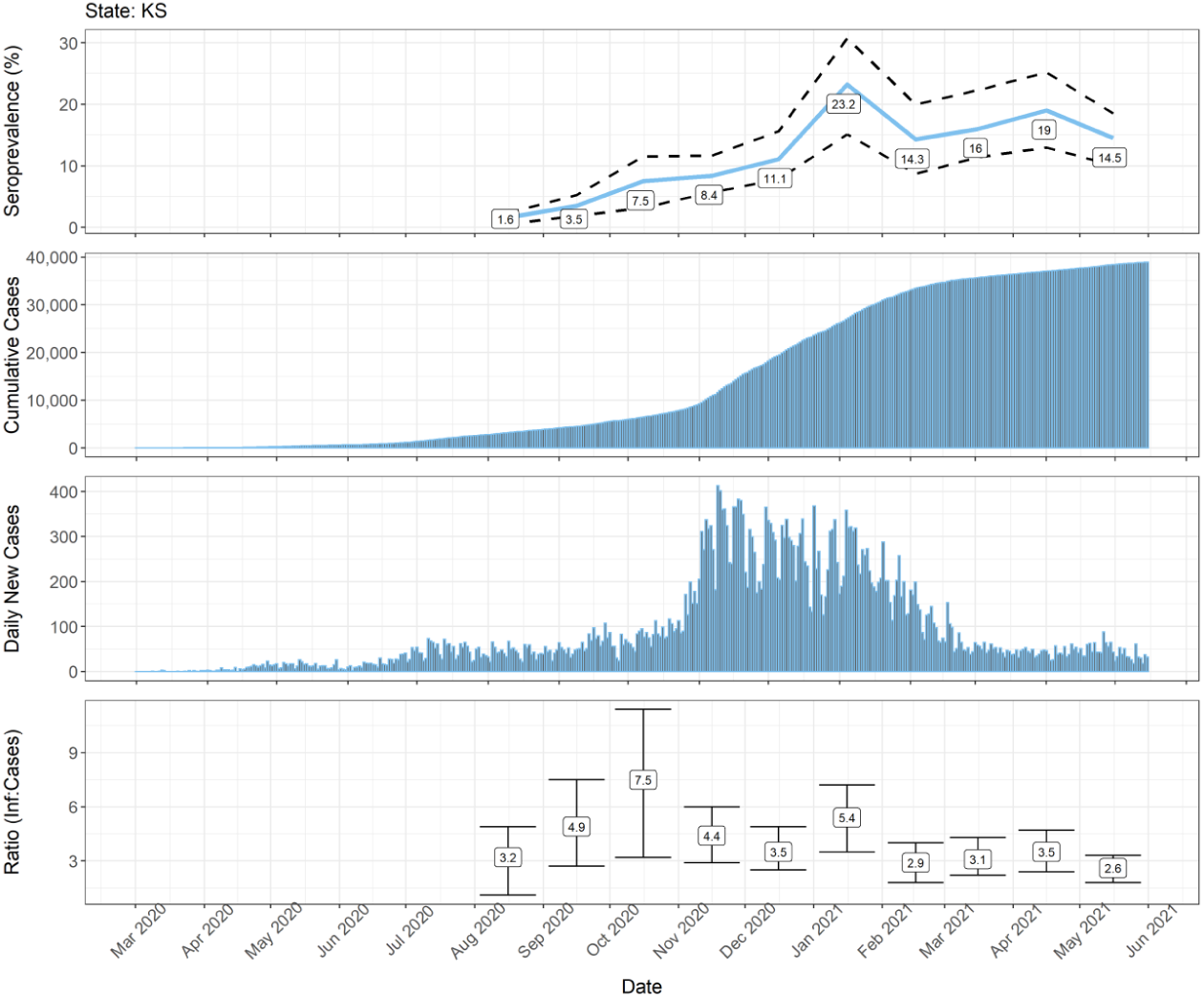
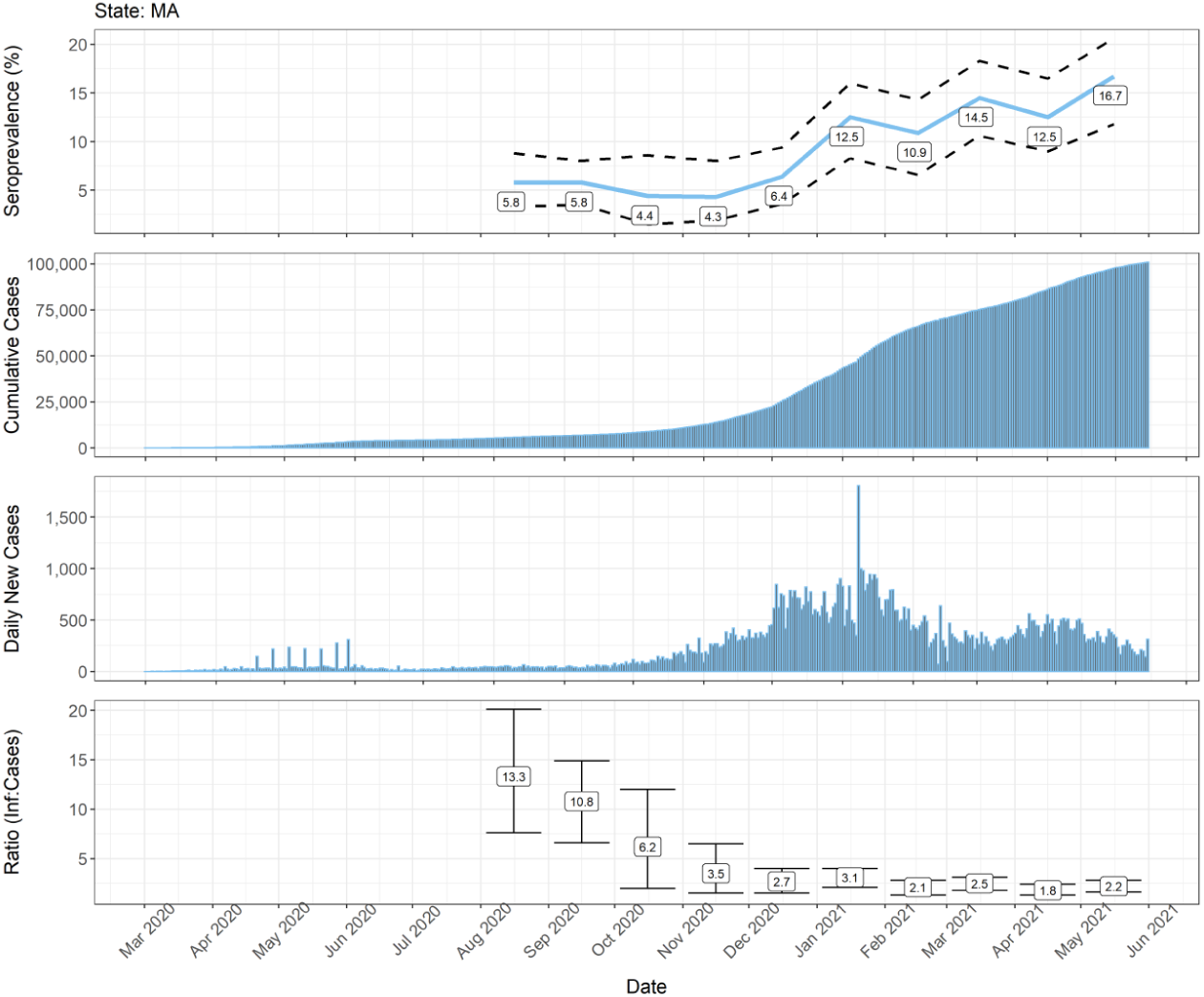
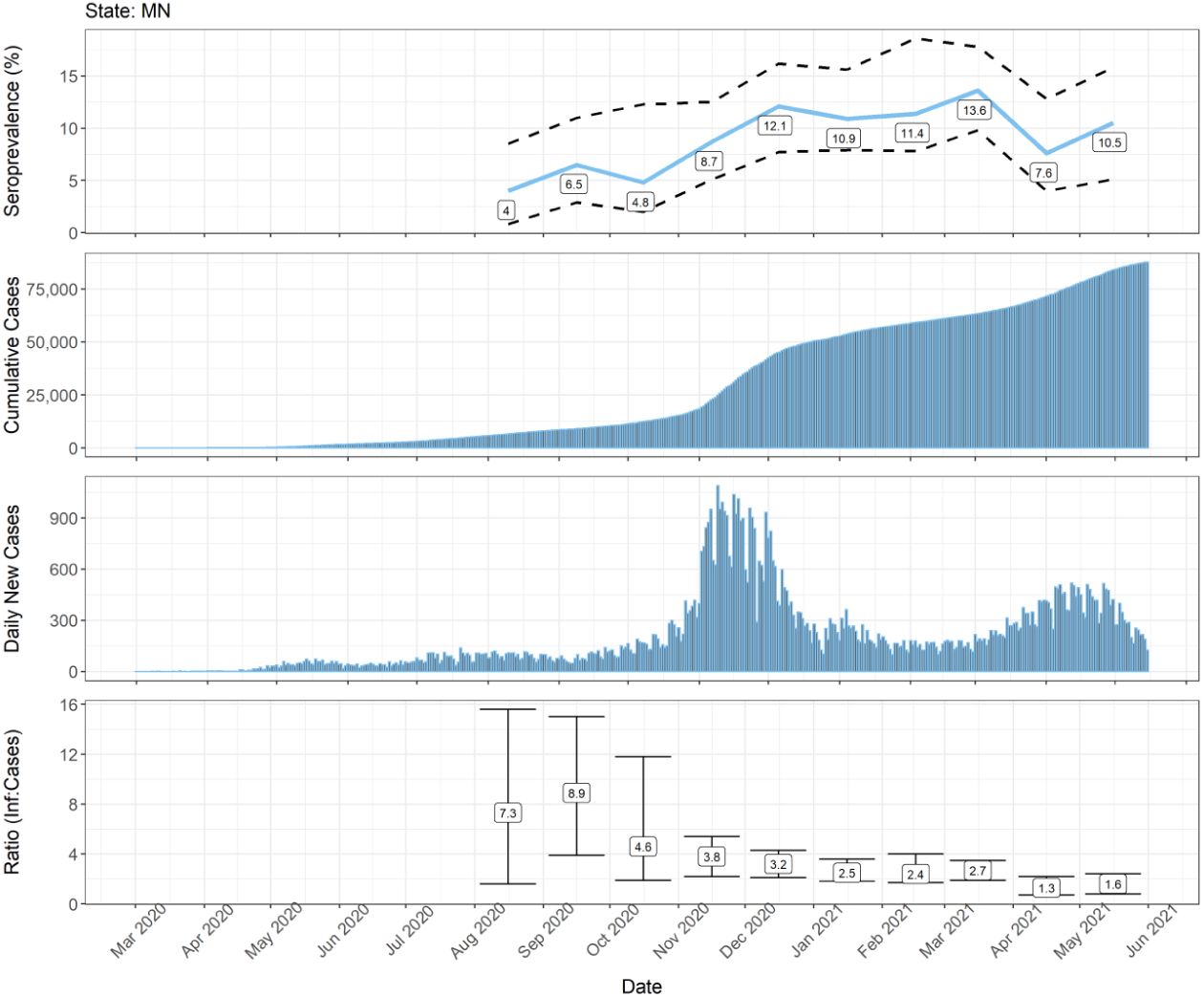
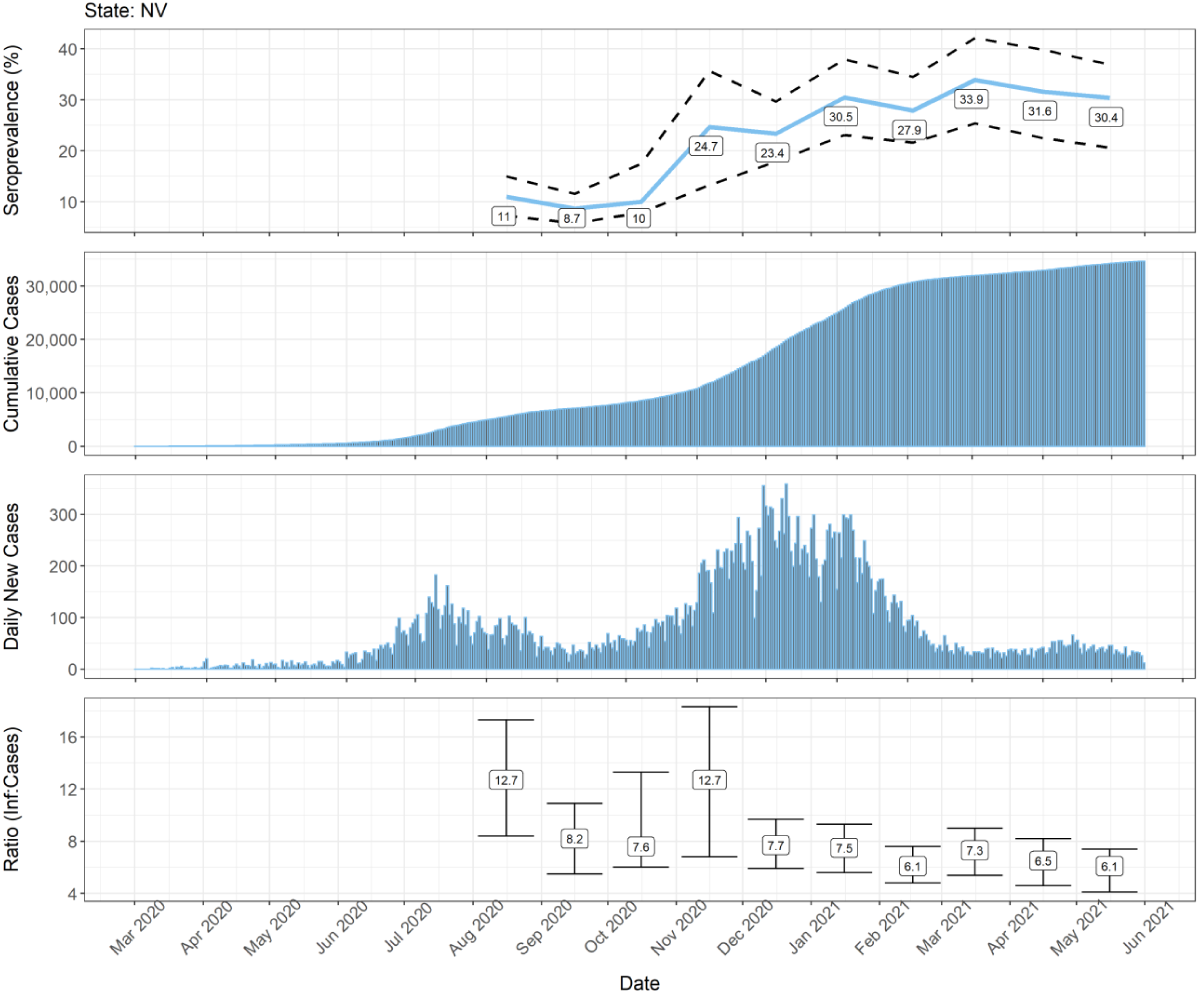
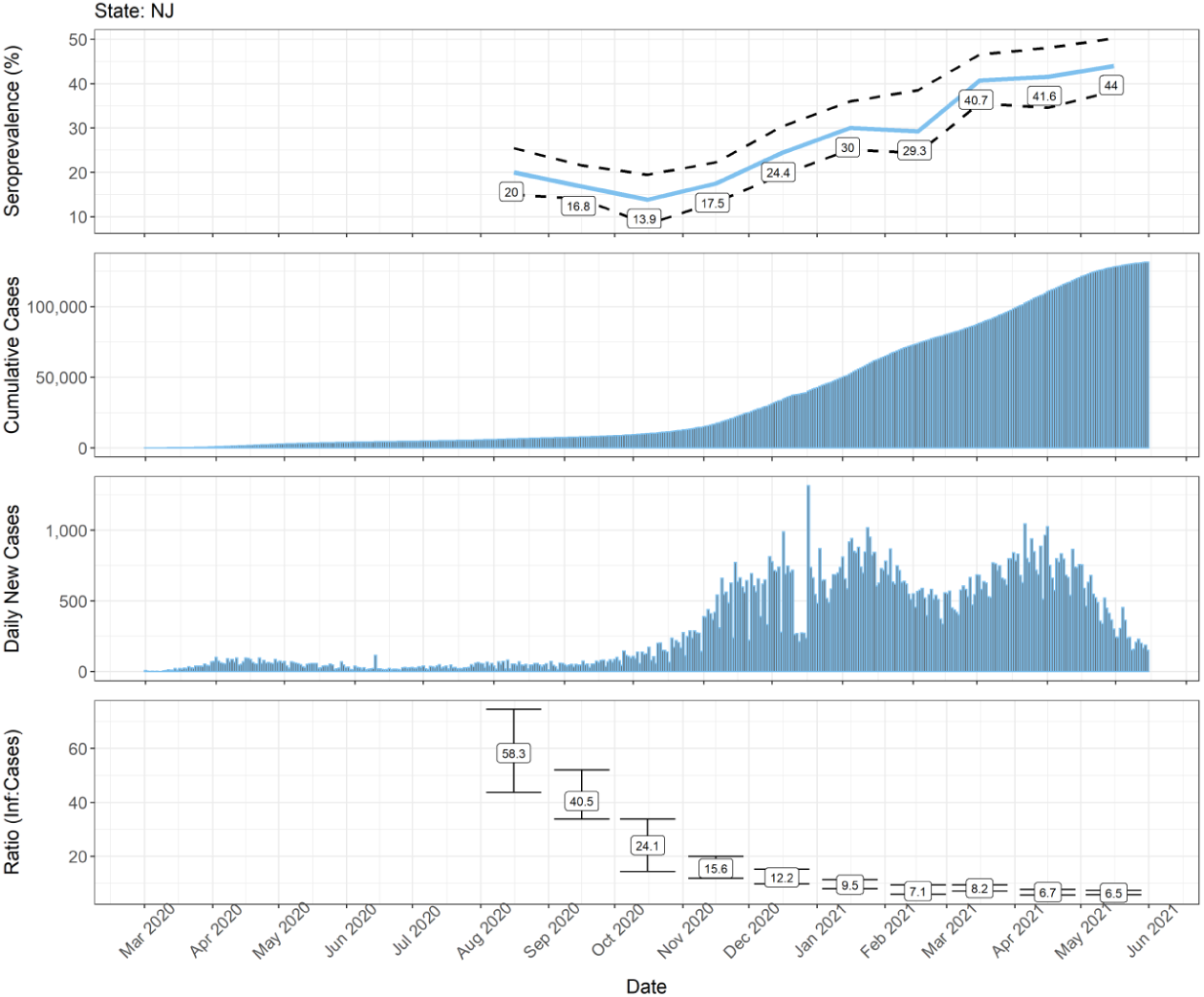
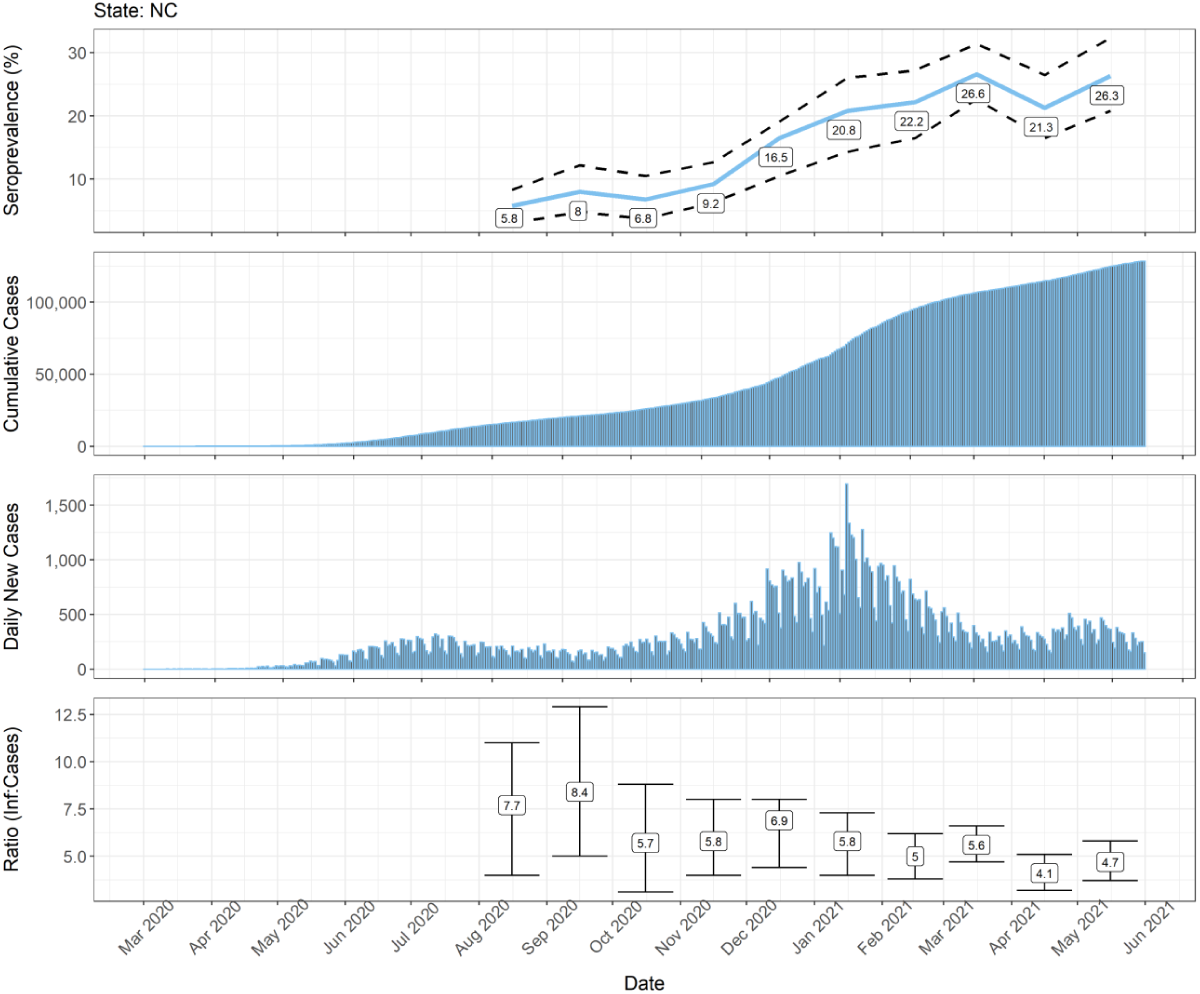
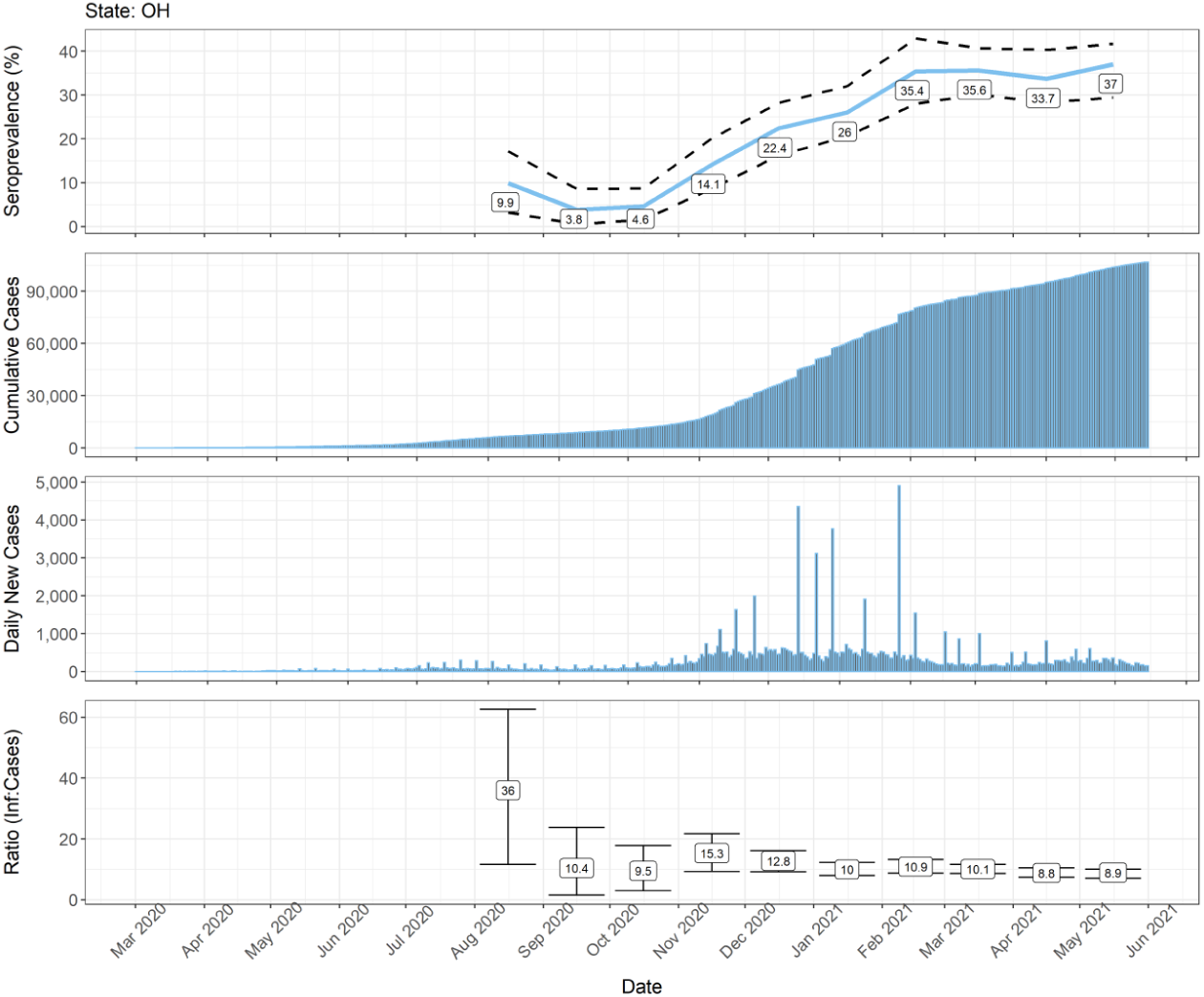
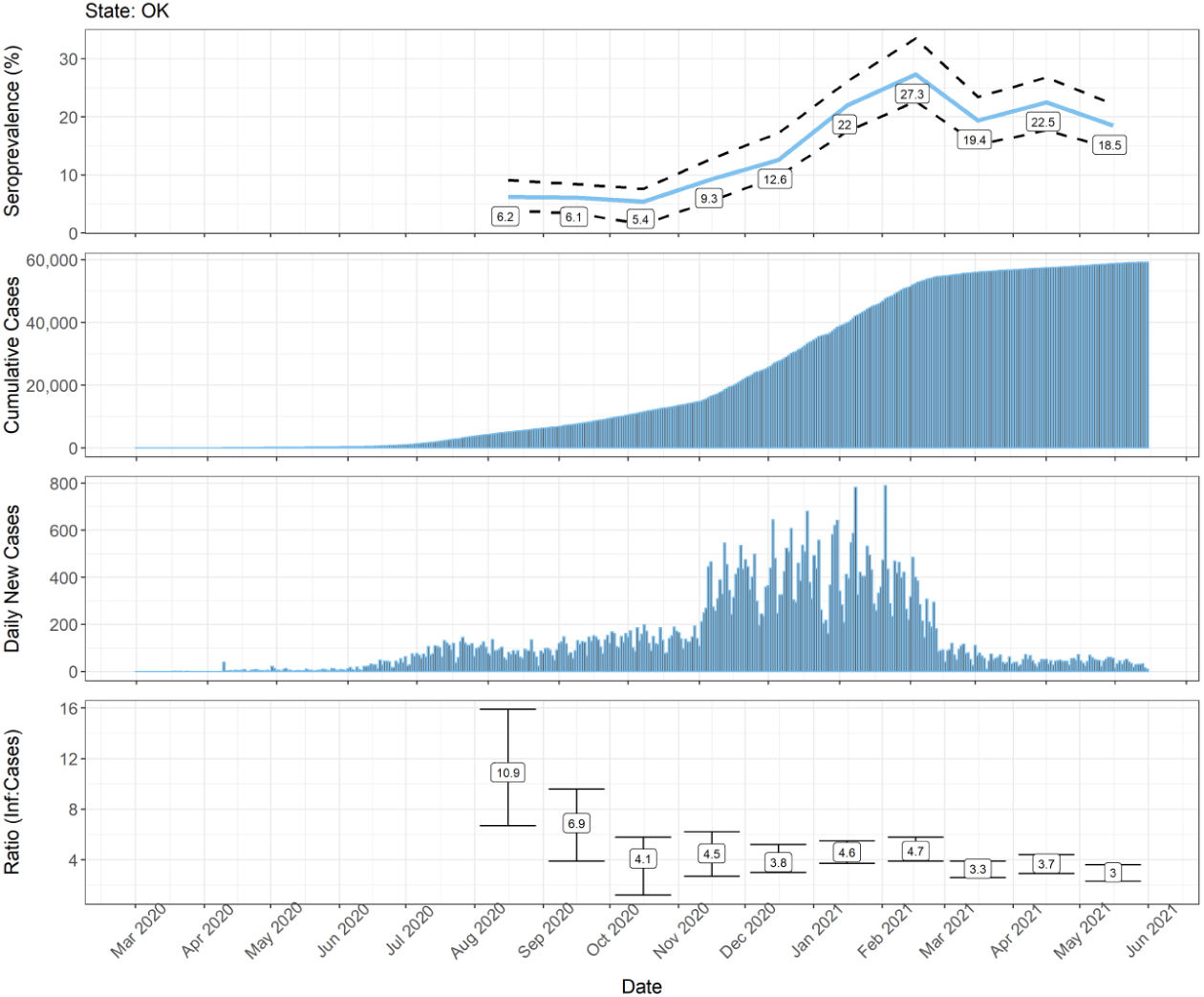
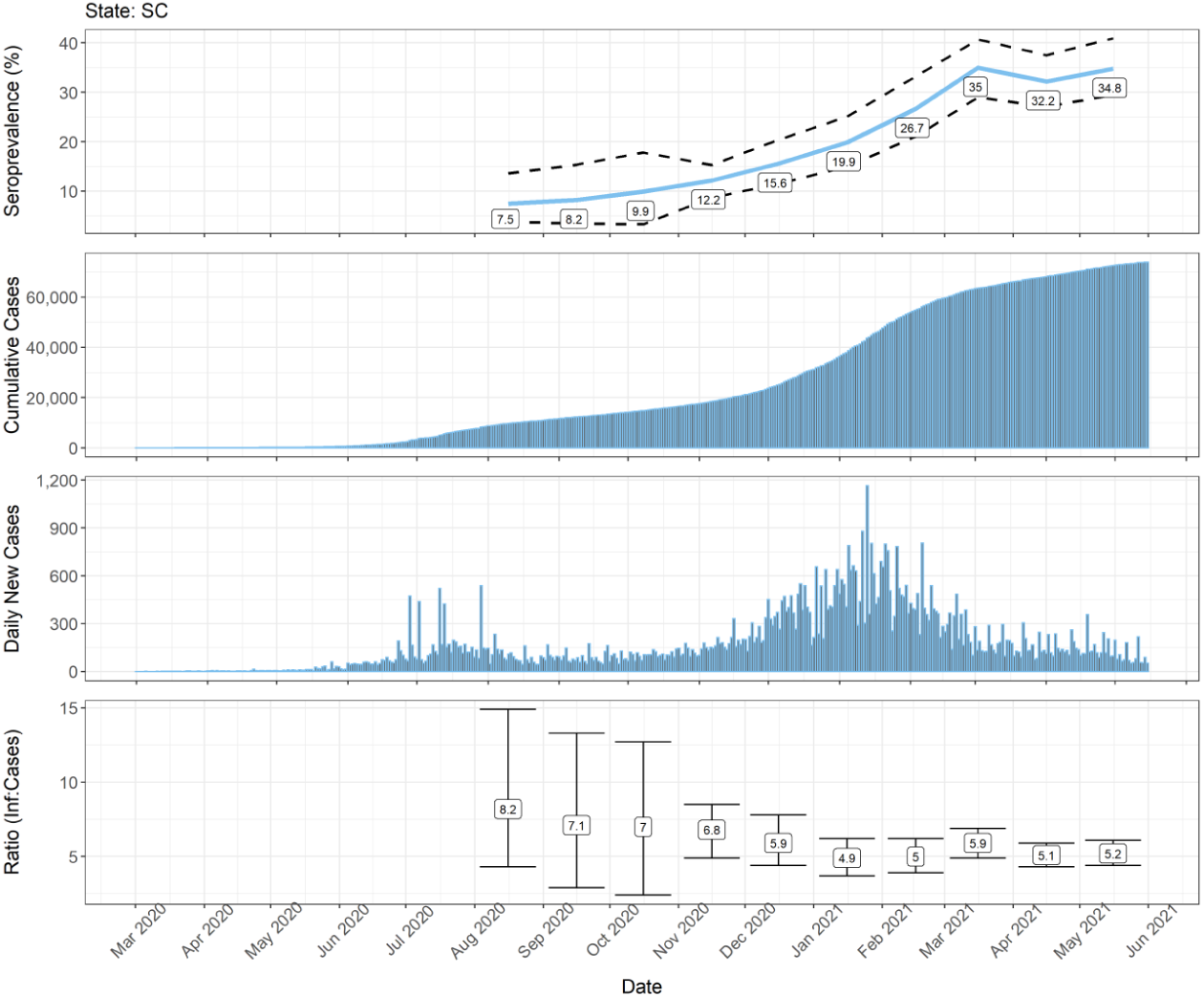
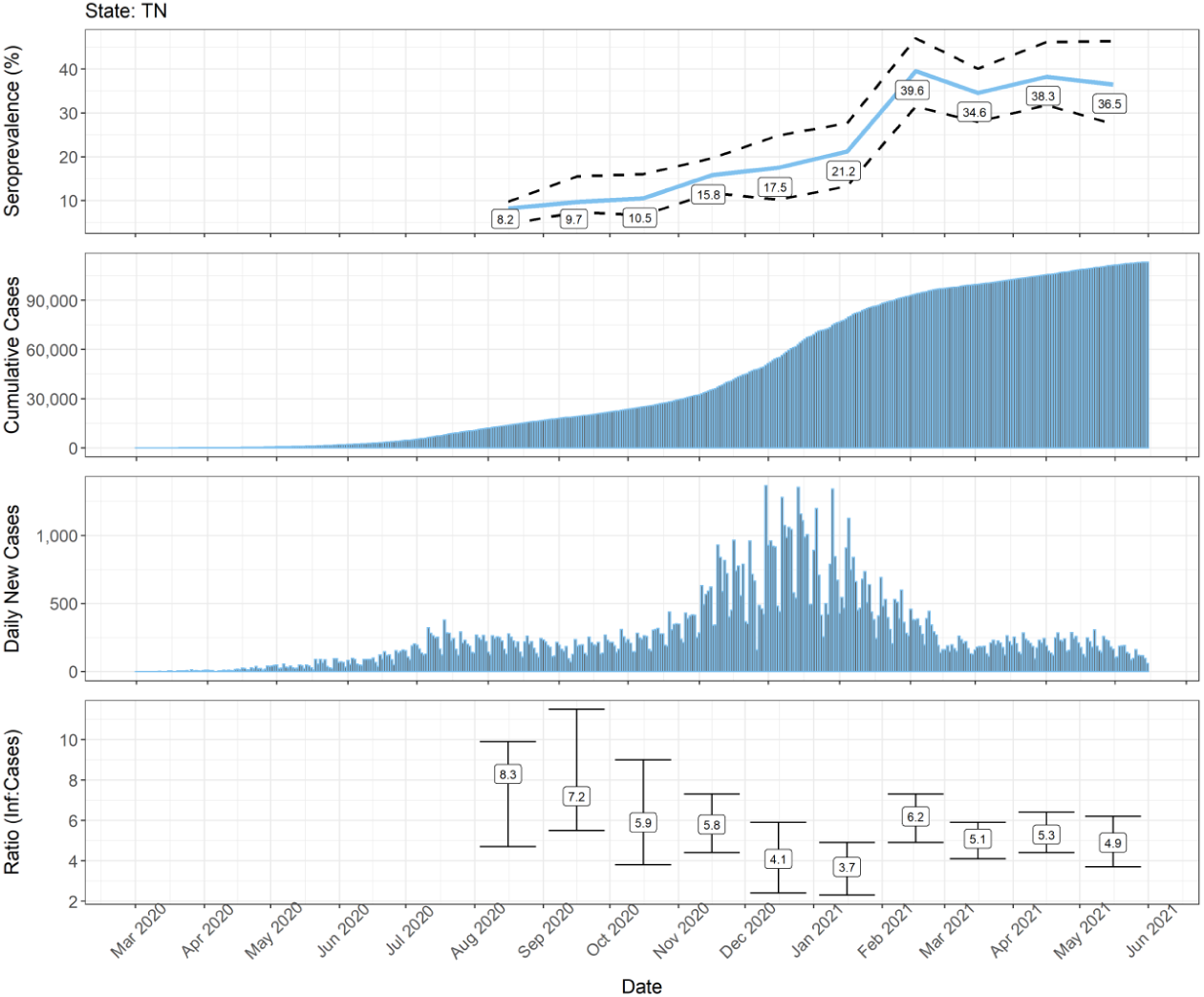
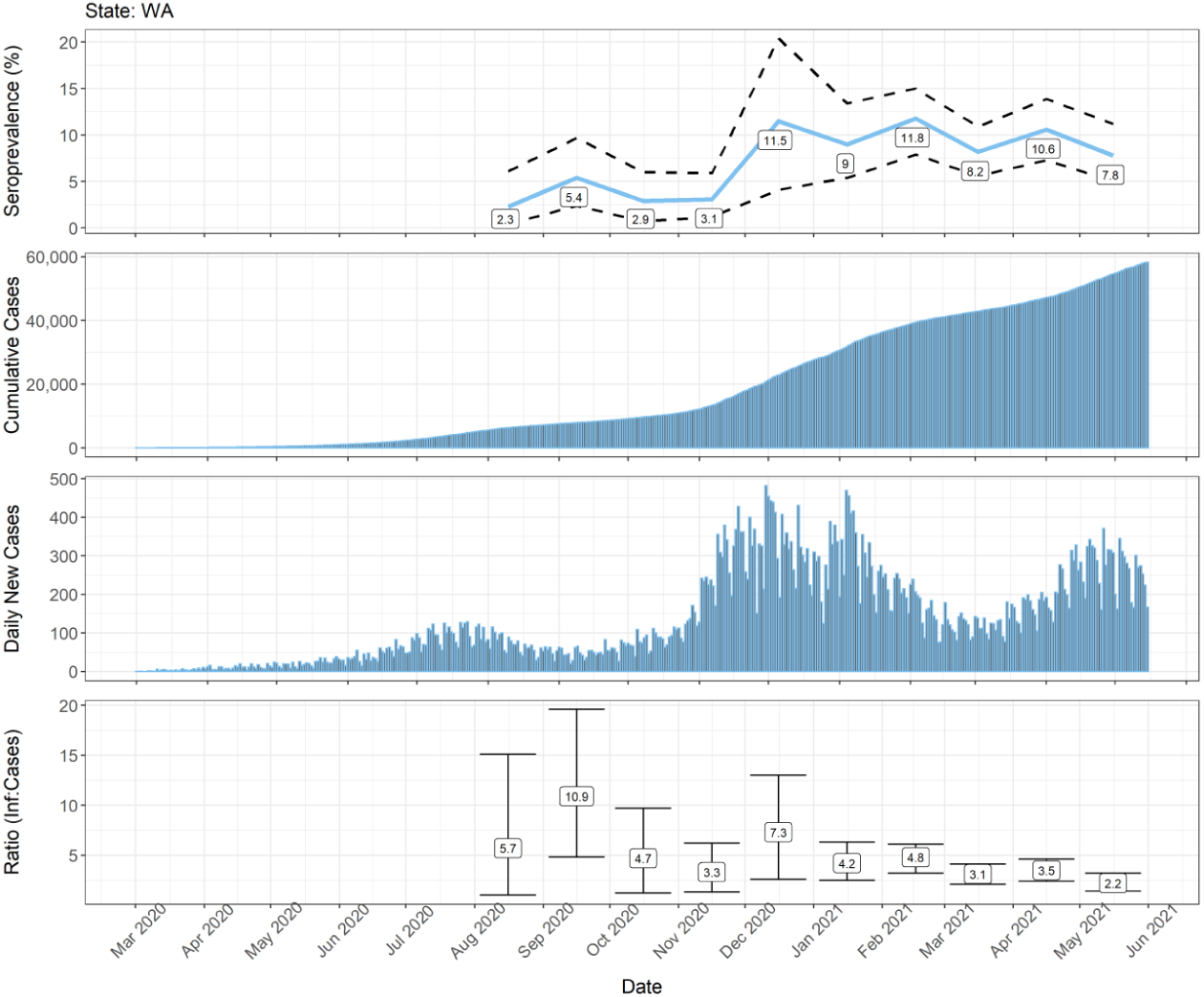
Estimated population-weighted SARS-CoV-2 seroprevalence by month of specimen collection, cumulative and daily reported COVID-19 confirmed and probable cases aged <18 years, and estimated SARS-CoV-2 infection to COVID-19 case ratio by state and calendar month, March 2020—May 2021.

